# Association between adverse childhood experiences and future cardiovascular disease risk in a population with Cardiovascular-Kidney-Metabolic syndrome stage 0-3: Depression symptom plays a mediating role

**DOI:** 10.1101/2025.10.22.25338593

**Authors:** Yankai Wang, Mingze Sun, Ji Gao, Jun Pu

## Abstract

**Background:** While adverse childhood experiences (ACEs) are established risk factors for cardiovascular disease (CVD), their association with CVD risk among individuals already at high risk due to Cardiovascular-Kidney-Metabolic (CKM) syndrome remains unclear. This study aimed to investigate this relationship and to evaluate the potential mediating role of depression symptoms.

**Methods:** This longitudinal study included 6,113 middle-aged and older adults with CKM syndrome (stages 0-3) from the China Health and Retirement Longitudinal Study (CHARLS). Cumulative ACE scores (total, intrafamilial, social) were derived from a life history survey. Incident CVD was determined during follow-up (2013-2018). Cox proportional hazards models assessed ACE-CVD associations, with restricted cubic splines evaluating dose-response relationships. Counterfactual-based mediation analysis quantified the contribution of depression symptoms (CES-D ≥10).

**Results:** During follow-up, 931 incident CVD cases occurred. A graded, positive association was observed between cumulative ACE exposure and CVD risk. In fully adjusted models, compared to the low ACE group (0-2), participants in medium (3-4) and high (≥5) ACE groups had significantly increased CVD risks (HR=1.22, 95% CI: 1.05-1.41; HR=1.29, 95% CI: 1.07-1.55, respectively). Each 1-point increase in the total ACE score was associated with a 7% elevated CVD risk (HR=1.07, 95% CI: 1.03-1.10). Intrafamilial ACEs demonstrated a stronger association (HR=1.08 per point, 95% CI: 1.04-1.13) than social ACEs. A nonlinear dose-response relationship was identified (P for nonlinearity <0.001). Depression symptoms significantly mediated 14.26% (95% CI: 4.56%-24.17%) of the total ACE-CVD association, with varying proportions observed for intrafamilial (15.92%) and social (32.47%) ACE subtypes.

**Conclusions:** Among Chinese adults with CKM syndrome stages 0-3, cumulative ACE exposure, particularly intrafamilial adversities, is independently associated with an increased risk of incident CVD in a dose-response manner. Depression symptoms partially mediate this association. Integrating ACE screening and addressing depression could enhance cardiovascular risk stratification and primary prevention in this high-risk population.

## Introduction

Cardiovascular-kidney-metabolic (CKM) syndrome is a systemic disorder driven by pathophysiological interactions among metabolic risk factors, chronic kidney disease, and the cardiovascular system.^1–3^ The escalating global prevalence of obesity and metabolic disturbances has coincided with a rising incidence of CKM syndrome, posing a substantial challenge to public health systems worldwide due to its associated burden.^4–6^ A recent Presidential Advisory from the American Heart Association underscores the critical importance of focusing research on populations with CKM stages 0 to 3 for the primary prevention of cardiovascular disease.^2^ Identifying modifiable risk factors during these preclinical stages is therefore essential to halt or delay disease progression and reduce the incidence of subsequent cardiovascular events. Adverse childhood experiences (ACEs), encompassing traumatic events such as emotional neglect, family violence, and parental mental illness during childhood, represent crucial early-life determinants of lifelong health.^7,8^ A substantial body of evidence indicates that ACEs are strongly associated not only with mental health disorders in adulthood but also with an elevated risk of various chronic physical diseases, particularly cardiovascular conditions.^9–11^ The underlying mechanisms are thought to involve chronic psychological stress induced by ACEs, which can lead to dysregulation of neuroendocrine, immune, and metabolic functions, thereby accelerating the development of atherosclerosis.^12–16^

Although the relationship between ACEs and cardiovascular health in the general population is well-established, it remains unclear whether ACEs constitute an independent risk factor for incident cardiovascular disease among individuals already at high risk due to CKM syndrome. Furthermore, the potential influence of ACEs on the progression through CKM stages itself is unknown. As an early-life stressor, ACEs may exert additive or interactive effects with this syndromic background, potentially amplifying cardiovascular risk. Depressive symptoms, a common sequela of ACEs and an established independent risk factor for cardiovascular disease,^17–20^ may thus serve as a significant mediating pathway linking early adversity to cardiovascular outcomes in this vulnerable population.

To address these knowledge gaps, this study leverages a large, nationally representative longitudinal sample from the China Health and Retirement Longitudinal Study (CHARLS). We aim to investigate whether ACEs, which assessed as cumulative scores, intrafamilial ACEs, and social ACEs, are independently associated with future cardiovascular disease risk among middle-aged and older Chinese adults with CKM stages 0–3. We further seek to characterize the dose-response relationship in this association and to evaluate the potential mediating role of depressive symptoms. Our findings clarify the role of childhood adversity in cardiovascular disease among individuals with CKM syndrome, providing a basis for early psychological intervention and primary cardiovascular prevention in this high-risk group.

## Methods

### Study population

This longitudinal study utilized data from the China Health and Retirement Longitudinal Study (CHARLS), a nationally representative cohort survey that includes adults aged 45 years and above from 150 counties and districts across 28 provinces in China. Following the baseline survey in 2011, CHARLS conducted follow-up surveys in 2013, 2015, and 2018, along with a life history survey in 2014 that recorded participants’ experiences since birth.

This study analyzed data from five CHARLS waves: wave 1 (2011), wave 2 (2013), wave 3 (2014), wave 4 (2015), and wave 5 (2018). Comprehensive information on cardiovascular–kidney–metabolic (CKM) syndrome and cardiovascular disease (CVD) was collected in waves 1, 2, 4, and 5, while information on depressive symptoms was collected in wave 1. Demographic characteristics and lifestyle variables were obtained at wave 1, and data on adverse childhood experiences (ACEs) were collected at wave 3.

Among the 17,705 participants enrolled in 2011, individuals aged <45 years or those with incomplete data on age, sex, marital status, education, smoking, drinking, or blood test data (n = 8304), as well as those with incomplete ACEs data (n = 1029), were excluded. To focus on participants at CKM stages 0–3 at baseline in 2011, individuals who had clinical cardiovascular disease at baseline (n = 2256) were also excluded. This resulted in 6,116 participants being included at baseline (Figure 1).

**Figure 1.**
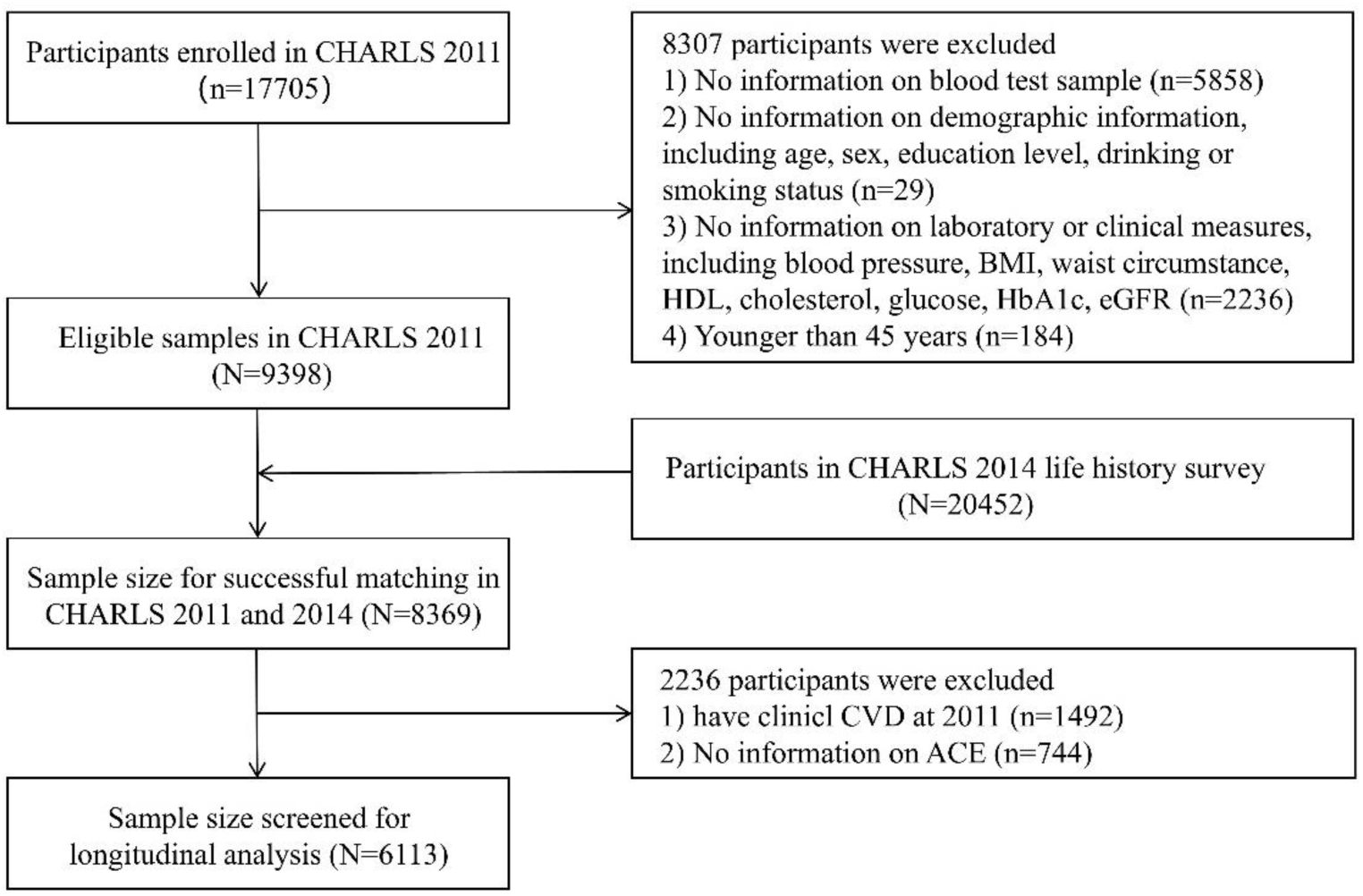
Flowchart of the study population

### Assessment of ACEs

Data on ACEs were derived from the 2014 Life History Survey. Participants retrospectively reported twelve types of early-life adversities covering both family and social domains. Intrafamilial ACEs included emotional neglect, family violence, parental separation or divorce, parental behavioral problems, parental mental illness, parental disability, parental death, physical abuse, and economic adversity. Social ACEs included bullying, loneliness, and unfriendly neighbors **(Table S1)**. Each ACE item was coded as present or absent (1/0) and summed to generate a cumulative ACE score ranging from 0 to 12.^21^ Participants were further categorized into three groups according to cumulative ACE exposure: 0–2 (Low), 3–4 (Medium), and ≥5 (High).

### CVD diagnosis

At each survey wave, participants were asked whether a physician had ever diagnosed them with any heart condition, including myocardial infarction, coronary heart disease, angina, heart failure, or other heart problems, or with stroke. Participants who reported either a heart condition or stroke were classified as having CVD. In this study, CVD status collected in the 2013, 2015, and 2018 waves was used to determine incident CVD events. All data were collected through standardized interviews under strict quality control procedures to ensure consistency and reliability.^22^

### Assessment of CKM syndrome across stage 0-3

CKM syndrome stages were determined according to the 2023 AHA Presidential Advisory criteria, adapted to CHARLS available biomarkers.^2^ Staging was performed at two time points (2011 and 2015) as only these survey waves contained the necessary laboratory measurements. Stage 0 included individuals with no evident CKM risk factors, defined by normal body mass index, waist circumference, blood pressure, fasting glucose, lipid profile, and kidney function, and without cardiovascular disease. Stage 1 represented excess or dysfunctional adiposity or impaired glucose metabolism. Stage 2 included participants with metabolic abnormalities or chronic kidney disease, characterized by hypertension, diabetes, dyslipidemia, or an estimated glomerular filtration rate below 60 mL/min/1.73 m². Stage 3 referred to the presence of subclinical cardiovascular disease or very high-risk CKD (eGFR < 30 mL/min/1.73 m²) without clinical CVD.^23^ The Chinese Modification of Diet in Renal Disease (C-MDRD) equation was used to calculate eGFR, and kidney function stages were classified according to KDIGO criteria. Participants with clinical CVD (CKM stage 4) were excluded at baseline **(Table S2-S5)**.

### Covariates

This study examines the impact of numerous covariates on the correlation of ACEs with CVD incidence within patients across CKM stages 0–3, based on prior research.^24^ Categorical variables included sex, marital status, education, smoking, and drinking status. Education was classified into three levels: college/university, secondary school, and primary school or lower. Smoking was defined based on the question “Smoke or Not,” with participants answering “yes” classified as smokers and those answering “no” as non-smokers. Drinking status was determined from the question “Did you drink any alcoholic beverages last year?” Participants reporting “drink more than once a month” or “drink but less than once a month” were categorized as drinkers, whereas those selecting “none of these” were considered non-drinkers. Continuous variables included age, systolic blood pressure (SBP), diastolic blood pressure (DBP), total cholesterol (TC), fasting blood glucose, high-density lipoprotein cholesterol (HDL-C), hemoglobin A1c (HbA1c), and estimated glomerular filtration rate (eGFR). SBP and DBP were calculated as the mean of three consecutive readings measured during the same visit. eGFR was calculated using the Chinese-adapted Modification of Diet in Renal Disease (MDRD) equation based on serum creatinine (SCr): eGFR = 175 × SCr⁻¹·²³⁴ × Age⁻⁰·¹⁷⁹ × 0.79 [if female].^25^ All covariates were collected according to standardized CHARLS protocols to ensure consistency and reliability across survey waves.

### Sensitivity analysis

We conducted comprehensive sensitivity analyses to evaluate the robustness of our findings and potential selection bias. First, we systematically assessed missing data patterns across all study variables and implemented multiple imputation using chained equations (MICE) with 5 imputed datasets and 10 iterations. Continuous variables were imputed via predictive mean matching, while categorical variables used appropriate methods: logistic regression for binary variables and polytomous regression for multinomial variables. The imputation model incorporated all analysis variables to ensure proper conditional distributions.

Following imputation, we validated the plausibility of imputed values by comparing distributions of key variables before and after imputation. This verification confirmed that imputed values maintained reasonable distributions consistent with observed data patterns, supporting the appropriateness of our imputation approach.

Finally, to assess potential selection bias from excluding participants with incomplete data, we compared baseline characteristics between the included analytical sample after multiple imputation (n=7,404) and excluded participants across demographic factors, clinical parameters, and behavioral factors using t-tests for continuous variables and chi-square tests for categorical variables.^26^

### Statistical analysis

Descriptive statistics were used to summarize baseline characteristics across ACE exposure groups, with continuous variables presented as means ± standard deviations or medians with interquartile ranges, and categorical variables as frequencies and percentages. Group differences were assessed using analysis of variance or chi-square tests as appropriate.

We constructed Cox proportional hazards models to assess the associations of ACE exposure with incident CVD (assessed during 2013, 2015, and 2018 follow-ups) and CKM progression (evaluated at 2015 follow-up) among participants free of clinical CVD at baseline. ACE exposure was analyzed in three forms: categorized into low, medium, and high groups; as a continuous total score; and as subtype scores (intrafamilial and social). Three hierarchical adjustment levels were applied: crude models, demographic-adjusted models (age, sex, BMI), and fully adjusted models (additionally including education, systolic blood pressure, smoking, and drinking status).

To assess potential nonlinear relationships, restricted cubic spline analyses with 3 knots were performed using the continuous ACE score, with model specifications parallel to the primary Cox models. Nonlinearity was tested via ANOVA comparisons between spline and linear specifications.

Subgroup analyses were conducted to examine effect modification by age, gender, marital status, education, smoking, drinking, and CKM stages. For each subgroup, separate Cox models were fitted adjusting for all covariates except the stratification variable, with interaction terms tested to assess effect modification. Multicollinearity was evaluated using generalized variance inflation factors (GVIF), with values <5 considered acceptable. Depression symptoms were assessed using the CES-D scale (cutoff ≥10).

Mediation analyses employed counterfactual-based methods via the regmedint package to decompose total effects of ACE (total, intrafamilial, and social) into natural direct and indirect effects through depression symptoms, with all models adjusted for the full covariate set. The proportion mediated was calculated as the natural indirect effect divided by the total effect. All analyses were performed using R version 4.4.1, with hazard ratios, odds ratios, and 95% confidence intervals reported. Statistical significance was set at two-sided P < 0.05.

## Results

### Participant characteristics

Table 1 summarizes the baseline characteristics of the 6,113 study participants overall and stratified by ACE groups (low, medium, high). The mean age of the cohort was 59.3 ± 9.3 years, with a nearly equal distribution of males (47%) and females (53%).

**Table 1.**
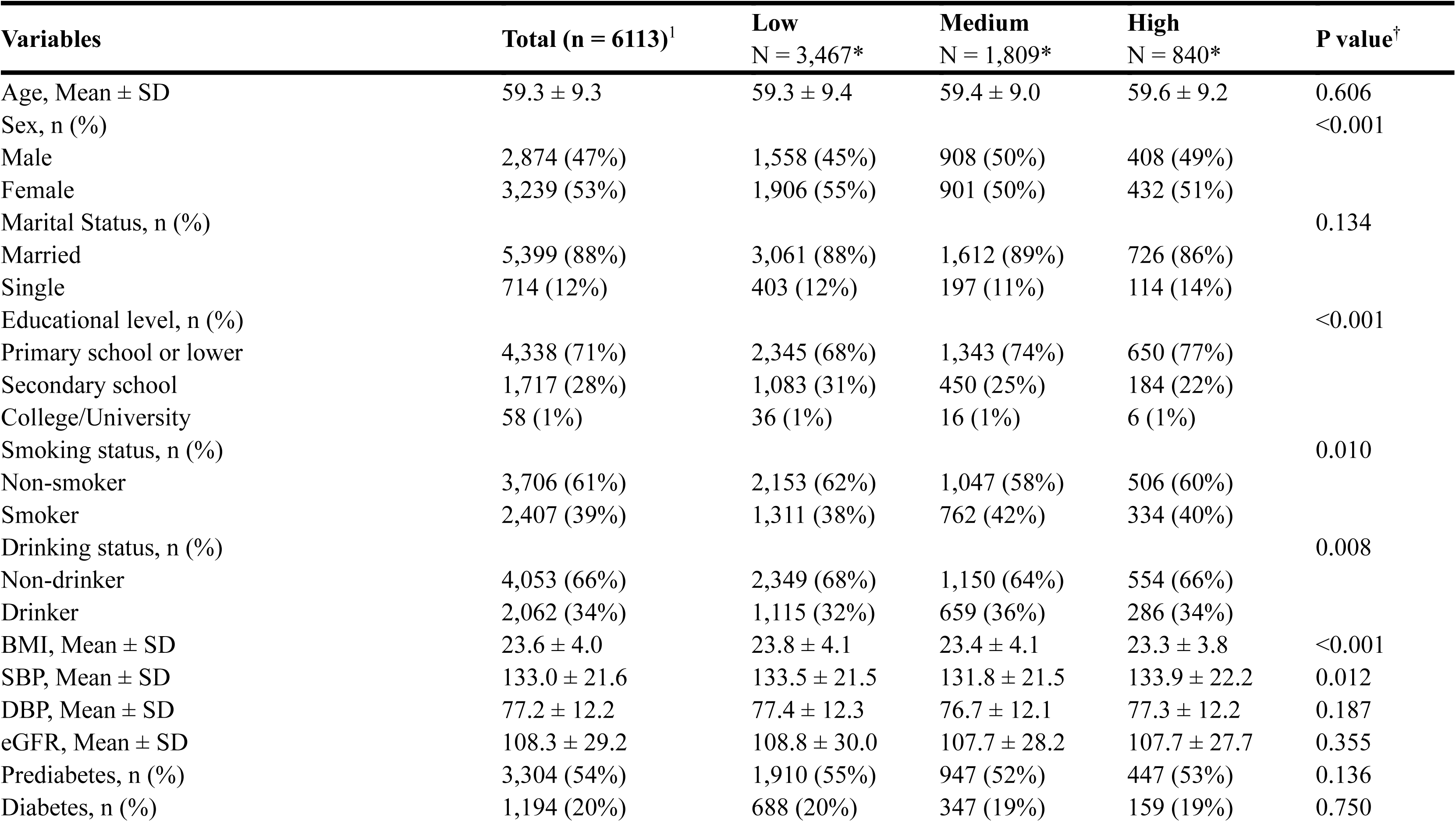
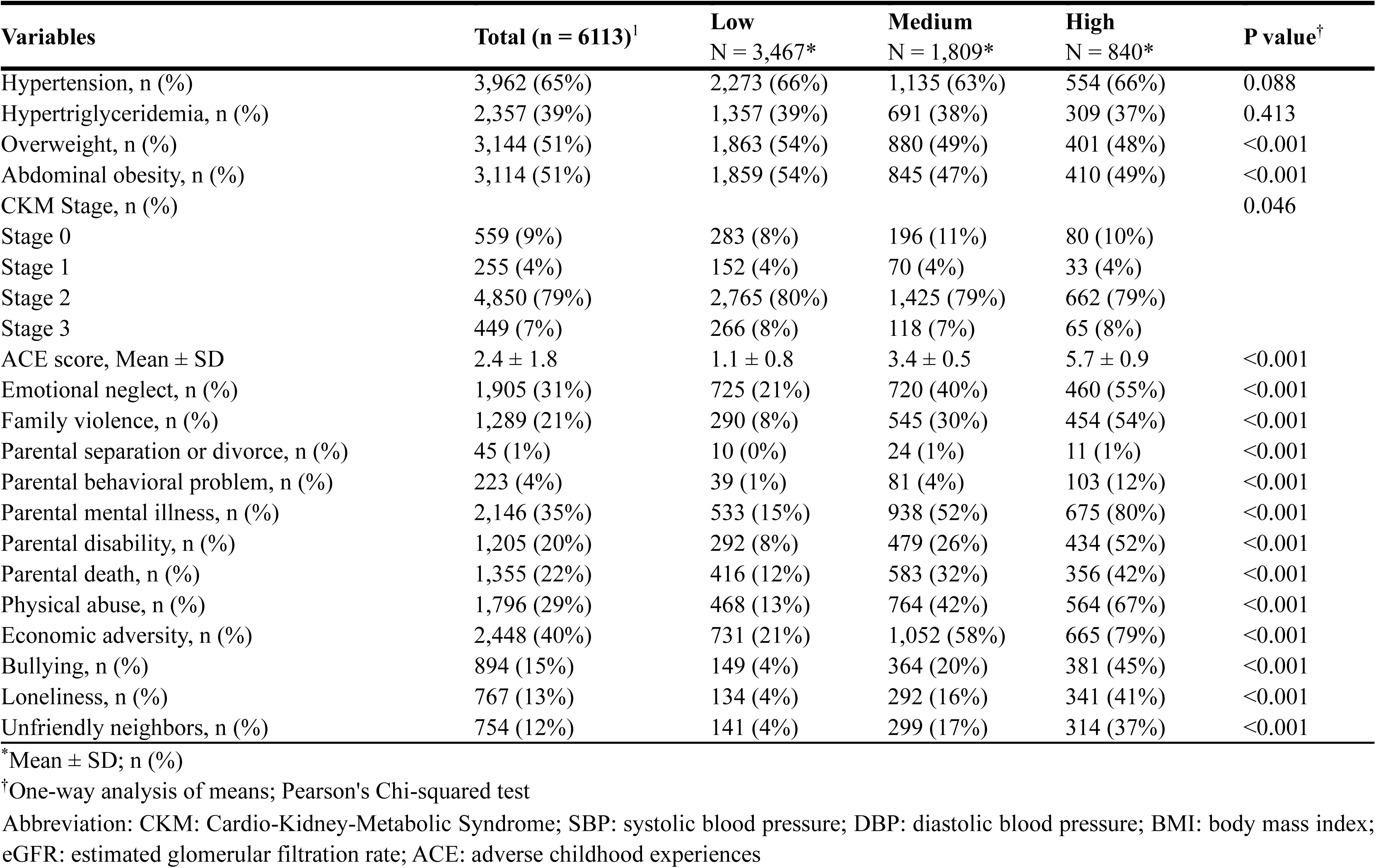
Baseline Characteristics of Participants according to ACE Groups.

Significant differences were observed across ACE groups for several key demographic and lifestyle factors. Participants with higher ACE exposure were more likely to have a lower educational attainment (p<0.001), with 77% in the high ACE group having primary school education or lower, compared to 68% in the low ACE group. Similarly, the distribution of sex, smoking, and drinking status also demonstrated significant disparities among the groups.

Regarding cardiometabolic risk profiles, significant gradients were noted across ACE groups for BMI, systolic blood pressure, and the prevalence of overweight and abdominal obesity. Specifically, the mean BMI was highest in the low ACE group (23.8 ± 4.1) and showed a decreasing trend with higher ACE exposure. The distribution of CKM stages also varied significantly by ACE group (p=0.046). As expected, the prevalence of all individual ACE components, such as emotional neglect, family violence, and economic adversity, exhibited a marked gradient increase from the low to the high ACE group (**Table 1**).

### Association between ACE and CVD incidence in a population with CKM syndrome stages 0–3

During the follow-up period, 931 incident CVD cases were documented among 6,113 participants. When ACE exposure was analyzed categorically, participants in the medium and high ACE groups showed significantly increased CVD risk compared to the low ACE group, with fully adjusted hazard ratios of 1.22 (95% CI: 1.05-1.41) and 1.29 (95% CI: 1.07-1.55), respectively. Analyzed as a continuous variable, each 1-point increase in the total ACE score was associated with a 7% elevated CVD risk (HR = 1.07, 95% CI: 1.03-1.10) in the fully adjusted model. When examining ACE subtypes, intrafamilial ACE demonstrated a significant association with CVD incidence (HR = 1.08 per point, 95% CI: 1.04-1.13), whereas social ACE did not reach statistical significance in any model specification.

For CKM syndrome progression, similar patterns were observed. Participants in medium and high ACE groups exhibited 14% (HR = 1.14, 95% CI: 1.03-1.27) and 27% (HR = 1.27, 95% CI: 1.11-1.45) increased risks of disease progression, respectively. Each additional total ACE point was associated with a 5% increased progression risk (HR = 1.05, 95% CI: 1.02-1.08). Intrafamilial ACE again showed a stronger association (HR = 1.07, 95% CI: 1.03-1.10) compared to social ACE, which was not significant in adjusted models (**Table 2**).

**Table 2.**
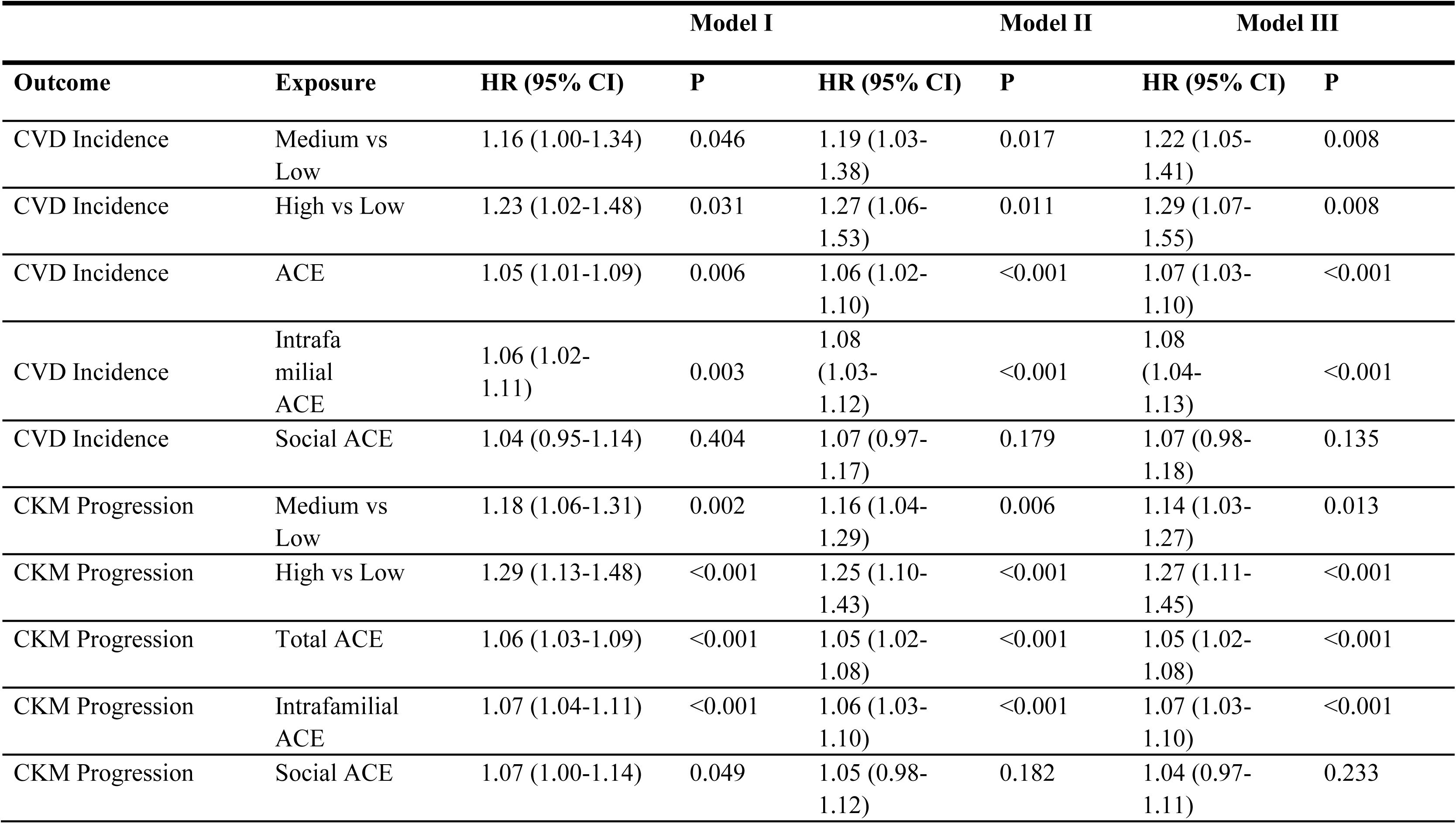
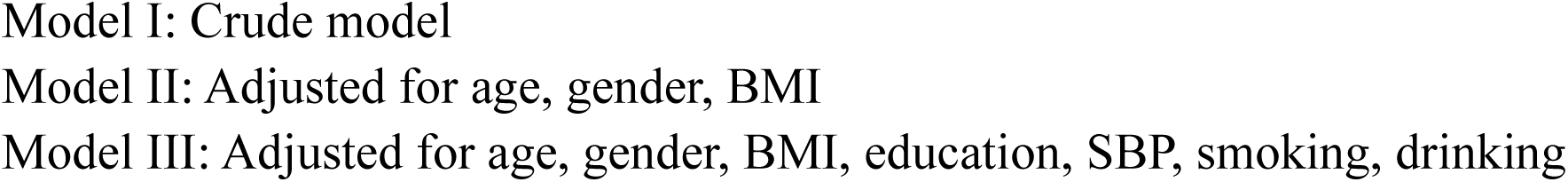
Association between ACE Measures and CVD Incidence and CKM Syndrome Progression in CKM Syndrome Stages 0-3.

| Outcome | Exposure | Model I |  | Model II |  | Model III |  |
| --- | --- | --- | --- | --- | --- | --- | --- |
|  |  | HR (95% CI) | P | HR (95% CI) | P | HR (95% CI) | P |
| CVD Incidence | Medium vs Low | 1.16 (1.00-1.34) | 0.046 | 1.19 (1.03-1.38) | 0.017 | 1.22 (1.05-1.41) | 0.008 |
| CVD Incidence | High vs Low | 1.23 (1.02-1.48) | 0.031 | 1.27 (1.06-1.53) | 0.011 | 1.29 (1.07-1.55) | 0.008 |
| CVD Incidence | ACE | 1.05 (1.01-1.09) | 0.006 | 1.06 (1.02-1.10) | <0.001 | 1.07 (1.03-1.10) | <0.001 |
| CVD Incidence | Intrafamilial ACE | 1.06 (1.02-1.11) | 0.003 | 1.08 (1.03-1.12) | <0.001 | 1.08 (1.04-1.13) | <0.001 |
| CVD Incidence | Social ACE | 1.04 (0.95-1.14) | 0.404 | 1.07 (0.97-1.17) | 0.179 | 1.07 (0.98-1.18) | 0.135 |
| CKM Progression | Medium vs Low | 1.18 (1.06-1.31) | 0.002 | 1.16 (1.04-1.29) | 0.006 | 1.14 (1.03-1.27) | 0.013 |
| CKM Progression | High vs Low | 1.29 (1.13-1.48) | <0.001 | 1.25 (1.10-1.43) | <0.001 | 1.27 (1.11-1.45) | <0.001 |
| CKM Progression | Total ACE | 1.06 (1.03-1.09) | <0.001 | 1.05 (1.02-1.08) | <0.001 | 1.05 (1.02-1.08) | <0.001 |
| CKM Progression | Intrafamilial ACE | 1.07 (1.04-1.11) | <0.001 | 1.06 (1.03-1.10) | <0.001 | 1.07 (1.03-1.10) | <0.001 |
| CKM Progression | Social ACE | 1.07 (1.00-1.14) | 0.049 | 1.05 (0.98-1.12) | 0.182 | 1.04 (0.97-1.11) | 0.233 |

Analysis of individual ACE components identified four specific adversities significantly associated with CVD risk in fully adjusted models: family violence (HR = 1.21, 95% CI: 1.05-1.39), parental mental illness (HR = 1.15, 95% CI: 1.02-1.30), parental disability (HR = 1.36, 95% CI: 1.18-1.56), and bullying (HR = 1.18, 95% CI: 1.01-1.39) **(Table S7)**.

Subgroup analyses demonstrated consistent positive associations between the continuous ACE score and CVD incidence across all predefined subgroups, including age, gender, marital status, education level, smoking and drinking status, and CKM stages **(Table S6)**. No significant interaction effects were observed, indicating the robustness of the ACE-CVD association across diverse participant characteristics.

Restricted cubic spline analyses revealed a significant nonlinear relationship between cumulative ACE exposure and CVD risk (P for nonlinearity < 0.001), with hazard ratios of 0.89 (95% CI: 0.83-0.96) at the 25th ACE percentile and 1.12 (95% CI: 1.03-1.21) at the 75th percentile relative to the median ACE score, suggesting an accelerated elevation in CVD risk at higher levels of cumulative ACE exposure (**Figure 2, Table S9)**.

**Figure 2.**
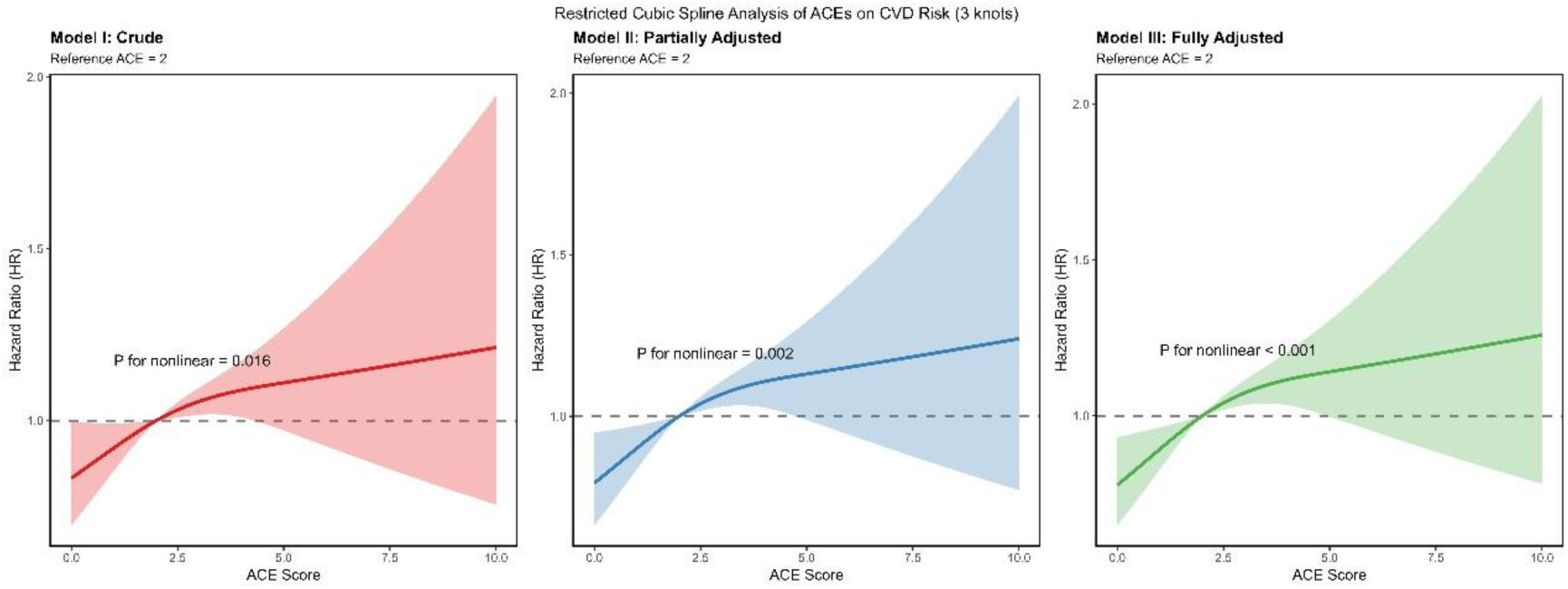
Restricted cubic spline analysis (RCS) of the association between cumulative ACE score and incident CVD risk in adults with CKM syndrome.

### Sensitivity analysis

To assess the robustness of our primary findings, we conducted a series of sensitivity analyses. First, we evaluated the potential impact of missing data. As shown in Table S8, several variables had a missing proportion of approximately 15%, while other covariates had minimal missingness. After applying multiple imputation for these missing covariates, the association between ACE exposure and CVD incidence remained consistent with the primary analysis **(Table S10)**. In the fully adjusted model using multiply imputed data, the hazard ratios for the medium and high ACE groups were 1.24 (95% CI: 1.09–1.41) and 1.27 (95% CI: 1.06–1.51), respectively, and each 1-point increase in the ACE score remained significantly associated with a 7% increased risk of CVD (HR=1.07, 95% CI: 1.03–1.10).

Second, we examined multicollinearity among all covariates included in the multivariable models. The variance inflation factors (VIFs) for all variables were well below the commonly accepted threshold of 5 (with the highest Generalized VIF being 2.89 for SBP), indicating that multicollinearity was not a substantial concern in our models **(Table S11)**.

Finally, to investigate potential selection bias, we compared the baseline characteristics of participants included in the final analytical sample after multiple imputation (n = 7,404) with those who were excluded (n = 4,443), a group primarily attributed to unsuccessful matching between CHARLS 2011 and 2014 waves, presence of clinical CVD at baseline in 2011, or lack of information on ACEs. While statistically significant differences were observed in gender, BMI, SBP, smoking and drinking status, and marital status **(Table S12)**, the absolute differences were small, suggesting that the exclusion of participants with missing data is unlikely to have substantially biased the observed associations.

### Mediation analyses for the associations of total ACE, intrafamilial ACE, and social ACE with CVD, mediated by depression symptoms

Mediation analyses were performed to examine whether depression symptoms mediated the association between ACE exposure and CVD incidence. As illustrated, depression symptoms significantly mediated the relationship between total ACE score and CVD (**Figure 3A)**. The natural indirect effect (NIE) was 1.013 (95% CI: 1.005, 1.021; P = 0.002), accounting for 14.26% (95% CI: 4.56%, 24.17%; P = 0.005) of the total effect. The natural direct effect (NDE) of total ACE on CVD remained statistically significant (1.065; 95% CI: 1.025, 1.106; P = 0.001). When examining ACE subtypes, depression symptoms also served as a significant mediator in the association between intrafamilial ACE and CVD, with a proportion mediated of 15.92% (95% CI: 4.76%, 27.08%; P = 0.005) (**Figure 3B)**. For social ACE, although the total effect on CVD was not statistically significant (1.099; 95% CI: 0.998, 1.213; P = 0.055), depression symptoms demonstrated a substantial mediating role, explaining 32.47% (95% CI: 14.25%, 69.23%; P = 0.003) of the association through a significant indirect effect (NIE: 1.030; 95% CI: 1.014, 1.047; P < 0.001) (**Figure 3C)**.

**Figure 3.**
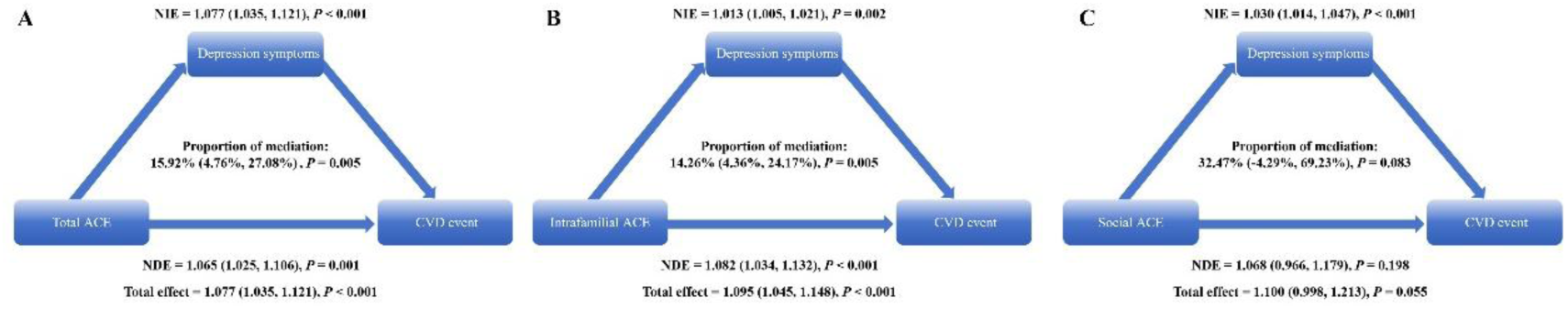
Mediation analysis of depressive symptoms on the association between total (A), intrafamilial (B), and social (C) ACEs and incident CVD risk.

These findings indicate that depression symptoms partially mediate the relationship between ACE exposure and CVD risk, with varying magnitudes of mediation across different ACE subtypes.

## Discussion

This national cohort study provides novel evidence linking adverse childhood experiences to cardiovascular disease risk among middle-aged and older Chinese adults with cardio-kidney-metabolic syndrome stages 0-3. We observed a graded association between cumulative ACE exposure and incident CVD, with each additional ACE increasing CVD risk. Restricted cubic spline analysis revealed a significant nonlinear relationship, marked by a sharp rise in risk at low ACE levels that plateaued with higher scores. When examining ACE subtypes, intrafamilial adversities demonstrated stronger associations with CVD than social adversities. A key finding was that depressive symptoms mediated approximately 15% of this relationship, suggesting mental health pathways substantially contribute to the ACE-CVD link in this population.

Our study confirms a significant association between ACEs and CVD risk in middle-aged and older adults, a finding consistent with most life-course epidemiological studies worldwide. Previous large-scale cohort studies have reported that cumulative ACE exposure is an independent risk factor for cardiovascular health in adulthood.^7,27,28^ Importantly, our restricted cubic spline analysis revealed a significant nonlinear dose-response relationship between ACEs and CVD risk. Unlike some studies reporting simple linear or J-shaped relationships,^7,29^ our analysis showed that the rate of increase in CVD risk gradually attenuated with higher ACE scores.

Furthermore, analysis of ACE subtypes indicated that intrafamilial ACEs, such as family violence and parental mental illness, were the primary drivers of CVD risk, whereas the independent association of social ACEs was relatively weaker. This observation aligns with research suggesting that intrafamilial adversities, due to their earlier onset, longer duration, and disruption of core attachment relationships, may exert more profound long-term health effects.^30^ Conversely, some evidence suggests that exposure to community threat or poverty may paradoxically correlate with fewer behavioral problems, potentially indicating adaptive coping.^31^ Nevertheless, other evidence has documented significant associations between social adversities, such as bullying, and cardiovascular risk markers.^32,33^ These inconsistencies may stem from variations in the definition, measurement, and exposure levels of social adversities across different cultural contexts, warranting further clarification in future research.

Our mediation analysis identified depressive symptoms as one pathway through which ACEs influence cardiovascular health. This finding is consistent with research showing that early life stress can alter brain development and stress response systems,^34^ increasing vulnerability to mood disorders. Depression, in turn, contributes to CVD risk through multiple channels, including adverse health behaviors^35–37^ and direct physiological effects on cardiovascular function.^38,39^

The biological processes underlying these relationships likely involve interrelated systems. Early adversity can trigger persistent dysregulation of stress response systems, primarily the hypothalamic-pituitary-adrenal (HPA) axis,^40,41^ leading to chronic inflammation^42,43^ that promotes atherosclerosis.^44^ These processes may converge to impair metabolic function and vascular integrity,^45,46^ particularly within the context of CKM syndrome where insulin resistance and endothelial dysfunction are hallmark features.^47,48^ Our findings position depressive symptoms within this complex network of biological and behavioral mechanisms linking childhood experiences to cardiovascular outcomes.

The consistent association between ACEs and CVD across all examined subgroups strengthens the evidence for ACEs as a fundamental cardiovascular risk factor. The absence of significant effect modification suggests that the detrimental impact of childhood adversity transcends conventional demographic and lifestyle categories.

Notably, the strongest point estimate emerged among participants with CKM stage 2, suggesting that ACEs may interact with established metabolic risk factors to amplify CVD vulnerability. The lack of significant associations in CKM stages 0 and 3 is primarily attributable to sparse data in these subgroups. Potential biological explanations, such as a longer latent period in stage 0 or masking by high baseline risk in stage 3, remain speculative without adequate sample sizes.

Methodological strengths of our study include the prospective design, nationally representative sampling, comprehensive assessment of ACEs and CKM staging, and advanced analytical approaches addressing mediation and nonlinearity. However, several limitations warrant consideration. Retrospective ACE assessment introduces potential recall bias, though this method remains standard in life-course epidemiology. Despite extensive covariate adjustment, residual confounding remains possible. Self-reported CVD outcomes, while practical for large cohorts, may lack the precision of clinical adjudication.

Clinically, our findings support integrating childhood adversity assessment into cardiovascular risk stratification for CKM patients. Identifying individuals with high ACE exposure could enable targeted screening for depression and early intervention to mitigate CVD risk. From a public health perspective, our results reinforce the importance of policies and programs that prevent childhood adversity and support healthy development. Future research should prioritize prospective studies with objective outcome measures, investigation of additional biological mechanisms beyond depression, and development of integrated interventions that address both psychological and cardiovascular health in ACE-affected populations.

## Conclusion

This study establishes a dose-response relationship between ACEs, particularly intrafamilial adversities, and increased cardiovascular disease risk in adults with CKM syndrome stages 0-3. Depression symptoms were identified as a significant mediator, explaining approximately 14% of this association. These findings advocate for the integration of ACE screening and depression management into cardiovascular risk stratification and primary prevention for this high-risk population. Ultimately, addressing childhood adversity and its psychological sequelae is crucial to mitigating long-term cardiovascular burdens.

## Data Availability

The data that support the findings of this study are from the China Health and Retirement Longitudinal Study (CHARLS). Researchers can obtain the data by applying through the official CHARLS website: http://charls.pku.edu.cn.

## Acknowledgments

This article uses data from CHARLS. The authors thank all the members of CHARLS for their time and efforts in the CHARLS project. All authors made significant contributions to the conception of the protocol, or the acquisition, analysis, or interpretation of the data. Yankai Wang and Mingze Sun drafted the manuscript. Jun Pu critically revised the manuscript. All authors have agreed to assume full responsibility for the integrity and accuracy of the manuscript and have reviewed and approved the final version for publication.

## Sources of Funding

This work was supported by the Noncommunicable Chronic Diseases-National Science and Technology Major Project (2023ZD0503204); the National Natural Science Foundation of China (82421001, 823B2005, 824B1015, 32470686 and 82230014); Shanghai Municipal Health Commission (2022JC013, 2023ZZ02021, 24ZR1443900 and GWVI-11.1-26); Shanghai Municipal Education Commission (SHSMU-ZDCX20210700).

## Disclosures

None.

## Supplemental Material

Tables S1–S12

## Notes

### Competing Interest Statement

The authors have declared no competing interest.

### Clinical Trial

This study is an observational analysis of data from the China Health and Retirement Longitudinal Study (CHARLS) and does not constitute a clinical trial. According to the International Committee of Medical Journal Editors (ICMJE) guidelines, prospective registration is required for interventional clinical trials, but not for observational studies such as this one. Therefore, a trial ID is not applicable.

### Author Declarations

The China Health and Retirement Longitudinal Study (CHARLS) was approved by the Institutional Review Board (IRB) at Peking University. The approval for the baseline survey was IRB00001052-11015. All participants provided written informed consent. This secondary data analysis study utilized de-identified, publicly available data from CHARLS. According to the policies of our institution and the Common Rule, such analysis does not constitute human subjects research and is therefore exempt from additional IRB review.

## References

1. Ndumele CE, Neeland IJ, Tuttle KR, Chow SL, Mathew RO, Khan SS, Coresh J, Baker-Smith CM, Carnethon MR, Després JP, et al. A Synopsis of the Evidence for the Science and Clinical Management of Cardiovascular-Kidney-Metabolic (CKM) Syndrome: A Scientific Statement From the American Heart Association. Circulation. 2023;148:1636–1664. doi: 10.1161/cir.0000000000001186

2. Ndumele CE, Rangaswami J, Chow SL, Neeland IJ, Tuttle KR, Khan SS, Coresh J, Mathew RO, Baker-Smith CM, Carnethon MR, et al. Cardiovascular-Kidney-Metabolic Health: A Presidential Advisory From the American Heart Association. Circulation. 2023;148:1606–1635. doi: 10.1161/cir.0000000000001184

3. Khan SS, Coresh J, Pencina MJ, Ndumele CE, Rangaswami J, Chow SL, Palaniappan LP, Sperling LS, Virani SS, Ho JE, et al. Novel Prediction Equations for Absolute Risk Assessment of Total Cardiovascular Disease Incorporating Cardiovascular-Kidney-Metabolic Health: A Scientific Statement From the American Heart Association. Circulation. 2023;148:1982–2004. doi: 10.1161/cir.0000000000001191

4. Aggarwal R, Ostrominski JW, Vaduganathan M. Prevalence of Cardiovascular-Kidney-Metabolic Syndrome Stages in US Adults, 2011-2020. Jama. 2024;331:1858–1860. doi: 10.1001/jama.2024.6892

5. Zhu R, Wang R, He J, Wang L, Chen H, Niu X, Sun Y, Guan Y, Gong Y, Zhang L, et al. Prevalence of Cardiovascular-Kidney-Metabolic Syndrome Stages by Social Determinants of Health. JAMA Netw Open. 2024;7:e2445309. doi: 10.1001/jamanetworkopen.2024.45309

6. Claudel SE, Schmidt IM, Waikar SS, Verma A. Cumulative Incidence of Mortality Associated with Cardiovascular-Kidney-Metabolic (CKM) Syndrome. J Am Soc Nephrol. 2025;36:1343–1351. doi: 10.1681/asn.0000000637

7. Lin L, Wang HH, Lu C, Chen W, Guo VY. Adverse Childhood Experiences and Subsequent Chronic Diseases Among Middle-aged or Older Adults in China and Associations With Demographic and Socioeconomic Characteristics. JAMA Netw Open. 2021;4:e2130143. doi: 10.1001/jamanetworkopen.2021.30143

8. Senaratne DNS, Thakkar B, Smith BH, Hales TG, Marryat L, Colvin LA. The impact of adverse childhood experiences on multimorbidity: a systematic review and meta-analysis. BMC Med. 2024;22:315. doi: 10.1186/s12916-024-03505-w

9. Wang W, Liu Y, Yang Y, Jiang W, Ni Y, Han X, Lu C, Guo L. Adverse childhood and adulthood experiences and risk of new-onset cardiovascular disease with consideration of social support: a prospective cohort study. BMC Med. 2023;21:297. doi: 10.1186/s12916-023-03015-1

10. Qiao Y, Zhu D, Zhao M, Magnussen CG, Xi B. Adverse childhood experience, adopting a healthy lifestyle in adulthood, and risk of cardiovascular diseases. J Affect Disord. 2024;362:450–458. doi: 10.1016/j.jad.2024.07.023

11. Pierce JB, Kershaw KN, Kiefe CI, Jacobs DR, Jr., Sidney S, Merkin SS, Feinglass J. Association of Childhood Psychosocial Environment With 30-Year Cardiovascular Disease Incidence and Mortality in Middle Age. J Am Heart Assoc. 2020;9:e015326. doi: 10.1161/jaha.119.015326

12. Yang JZ, Kang CY, Yuan J, Zhang Y, Wei YJ, Xu L, Zhou F, Fan X. Effect of adverse childhood experiences on hypothalamic-pituitary-adrenal (HPA) axis function and antidepressant efficacy in untreated first episode patients with major depressive disorder. Psychoneuroendocrinology. 2021;134:105432. doi: 10.1016/j.psyneuen.2021.105432

13. Li C, Xiang S. Adverse Childhood Experiences, Inflammation, and Depressive Symptoms in Late Life: A Population-Based Study. J Gerontol B Psychol Sci Soc Sci. 2023;78:220–229. doi: 10.1093/geronb/gbac179

14. Kivimäki M, Bartolomucci A, Kawachi I. The multiple roles of life stress in metabolic disorders. Nat Rev Endocrinol. 2023;19:10–27. doi: 10.1038/s41574-022-00746-8

15. Barrett TJ, Corr EM, van Solingen C, Schlamp F, Brown EJ, Koelwyn GJ, Lee AH, Shanley LC, Spruill TM, Bozal F, et al. Chronic stress primes innate immune responses in mice and humans. Cell Rep. 2021;36:109595. doi: 10.1016/j.celrep.2021.109595

16. Wang Y, Ke D, Chen Y, Zhang C, Liu W, Chen L, Pu J. Decoding immune aging at single-cell resolution. Trends Immunol. 2025. doi: 10.1016/j.it.2025.09.001

17. Antoniou G, Lambourg E, Steele JD, Colvin LA. The effect of adverse childhood experiences on chronic pain and major depression in adulthood: a systematic review and meta-analysis. Br J Anaesth. 2023;130:729–746. doi: 10.1016/j.bja.2023.03.008

18. Li J, Sun Q, Zhang H, Li B, Zhang C, Zhao Y, Lu J. Depressive symptoms mediate associations of adverse childhood experiences and chronic lung diseases: A mediation effect analysis. J Affect Disord. 2024;345:342–348. doi: 10.1016/j.jad.2023.10.140

19. Krittanawong C, Maitra NS, Qadeer YK, Wang Z, Fogg S, Storch EA, Celano CM, Huffman JC, Jha M, Charney DS, et al. Association of Depression and Cardiovascular Disease. Am J Med. 2023;136:881–895. doi: 10.1016/j.amjmed.2023.04.036

20. Qiu W, Cai A, Li L, Feng Y. Association of depression trajectories and subsequent hypertension and cardiovascular disease: findings from the CHARLS cohort. J Hypertens. 2024;42:432–440. doi: 10.1097/hjh.0000000000003609

21. Zheng G, Fang Z, Zhou B, He F, Zhang H, Xiao H, Qin X, Sun L, Zhu H, Hao G, et al. Association between adverse childhood experiences and long-term blood pressure variability: Insights from a pooled analysis. J Affect Disord. 2025;384:118–124. doi: 10.1016/j.jad.2025.05.039

22. Zhao Y, Hu Y, Smith JP, Strauss J, Yang G. Cohort profile: the China Health and Retirement Longitudinal Study (CHARLS). Int J Epidemiol. 2014;43:61–68. doi: 10.1093/ije/dys203

23. Wang X, Wen P, Liao Y, Wu T, Zeng L, Huang Y, Song X, Xiong Z, Deng L, Li D, et al. Association of atherogenic index of plasma and its modified indices with stroke risk in individuals with cardiovascular-kidney-metabolic syndrome stages 0-3: a longitudinal analysis based on CHARLS. Cardiovasc Diabetol. 2025;24:254. doi: 10.1186/s12933-025-02784-8

24. Tan MY, Zhang YJ, Zhu SX, Wu S, Zhang P, Gao M. The prognostic significance of stress hyperglycemia ratio in evaluating all-cause and cardiovascular mortality risk among individuals across stages 0-3 of cardiovascular-kidney-metabolic syndrome: evidence from two cohort studies. Cardiovasc Diabetol. 2025;24:137. doi: 10.1186/s12933-025-02689-6

25. Ma YC, Zuo L, Chen JH, Luo Q, Yu XQ, Li Y, Xu JS, Huang SM, Wang LN, Huang W, et al. Modified glomerular filtration rate estimating equation for Chinese patients with chronic kidney disease. J Am Soc Nephrol. 2006;17:2937–2944. doi: 10.1681/asn.2006040368

26. Wang X, Si H, Li Y, Yu J, Zhou W, Chen H, Wang C. Adverse childhood experiences and circadian syndrome in middle-aged and older adults: A multi-method analysis of a national sample. Maturitas. 2025;201:108695. doi: 10.1016/j.maturitas.2025.108695

27. Cui C, Liu L, Li H, Qi Y, Song J, Han N, Wang Z, Shang X, Sheng C, Balmer L, et al. Childhood Exposure to Interparental Physical Violence and Adult Cardiovascular Disease. JAMA Netw Open. 2024;7:e2451806. doi: 10.1001/jamanetworkopen.2024.51806

28. Hughes K, Bellis MA, Hardcastle KA, Sethi D, Butchart A, Mikton C, Jones L, Dunne MP. The effect of multiple adverse childhood experiences on health: a systematic review and meta-analysis. Lancet Public Health. 2017;2:e356–e366. doi: 10.1016/s2468-2667(17)30118-4

29. Bethell C, Jones J, Gombojav N, Linkenbach J, Sege R. Positive Childhood Experiences and Adult Mental and Relational Health in a Statewide Sample: Associations Across Adverse Childhood Experiences Levels. JAMA Pediatr. 2019;173:e193007. doi: 10.1001/jamapediatrics.2019.3007

30. Wade R, Jr., Cronholm PF, Fein JA, Forke CM, Davis MB, Harkins-Schwarz M, Pachter LM, Bair-Merritt MH. Household and community-level Adverse Childhood Experiences and adult health outcomes in a diverse urban population. Child Abuse Negl. 2016;52:135–145. doi: 10.1016/j.chiabu.2015.11.021

31. Russell JD, Heyn SA, Peverill M, DiMaio S, Herringa RJ. Traumatic and Adverse Childhood Experiences and Developmental Differences in Psychiatric Risk. JAMA Psychiatry. 2025;82:66–74. doi: 10.1001/jamapsychiatry.2024.3231

32. Baldwin JR, Arseneault L, Caspi A, Fisher HL, Moffitt TE, Odgers CL, Pariante C, Ambler A, Dove R, Kepa A, et al. Childhood victimization and inflammation in young adulthood: A genetically sensitive cohort study. Brain Behav Immun. 2018;67:211–217. doi: 10.1016/j.bbi.2017.08.025

33. Lüscher TF. Forgotten cardiovascular risk factors: pregnancy complications and preterm birth, bullying, periodontal disease, and hypoxic burden. Eur Heart J. 2019;40:1093–1096. doi: 10.1093/eurheartj/ehz171

34. van der Weerd N, Mulders P, Vrijsen J, van Oort J, Collard R, van Eijndhoven P, Tendolkar I. Childhood adversity, stress reactivity, and structural brain measures in stress-related/neurodevelopmental disorders, and their comorbidity: A large transdiagnostic cross-sectional study. Hum Brain Mapp. 2024;45:e70025. doi: 10.1002/hbm.70025

35. Kiecolt-Glaser JK, Derry HM, Fagundes CP. Inflammation: depression fans the flames and feasts on the heat. Am J Psychiatry. 2015;172:1075–1091. doi: 10.1176/appi.ajp.2015.15020152

36. Fluharty M, Taylor AE, Grabski M, Munafò MR. The Association of Cigarette Smoking With Depression and Anxiety: A Systematic Review. Nicotine Tob Res. 2017;19:3–13. doi: 10.1093/ntr/ntw140

37. Swainson J, Reeson M, Malik U, Stefanuk I, Cummins M, Sivapalan S. Diet and depression: A systematic review of whole dietary interventions as treatment in patients with depression. J Affect Disord. 2023;327:270–278. doi: 10.1016/j.jad.2023.01.094

38. Zhang Z, Xu H, Zhang R, Yan Y, Ling X, Meng Y, Zhang X, Wang Y. Frailty and depressive symptoms in relation to cardiovascular disease risk in middle-aged and older adults. Nat Commun. 2025;16:6008. doi: 10.1038/s41467-025-61089-2

39. Harshfield EL, Pennells L, Schwartz JE, Willeit P, Kaptoge S, Bell S, Shaffer JA, Bolton T, Spackman S, Wassertheil-Smoller S, et al. Association Between Depressive Symptoms and Incident Cardiovascular Diseases. Jama. 2020;324:2396–2405. doi: 10.1001/jama.2020.23068

40. Herzberg MP, Gunnar MR. Early life stress and brain function: Activity and connectivity associated with processing emotion and reward. Neuroimage. 2020;209:116493. doi: 10.1016/j.neuroimage.2019.116493

41. Heim C, Nemeroff CB. The role of childhood trauma in the neurobiology of mood and anxiety disorders: preclinical and clinical studies. Biol Psychiatry. 2001;49:1023–1039. doi: 10.1016/s0006-3223(01)01157-x

42. Cruz-Pereira JS, Rea K, Nolan YM, O’Leary OF, Dinan TG, Cryan JF. Depression’s Unholy Trinity: Dysregulated Stress, Immunity, and the Microbiome. Annu Rev Psychol. 2020;71:49–78. doi: 10.1146/annurev-psych-122216-011613

43. Michopoulos V, Powers A, Gillespie CF, Ressler KJ, Jovanovic T. Inflammation in Fear- and Anxiety-Based Disorders: PTSD, GAD, and Beyond. Neuropsychopharmacology. 2017;42:254–270. doi: 10.1038/npp.2016.146

44. Henein MY, Vancheri S, Longo G, Vancheri F. The Role of Inflammation in Cardiovascular Disease. Int J Mol Sci. 2022;23. doi: 10.3390/ijms232112906

45. Belkin S, Benthien J, Axt PN, Mohr T, Mortensen K, Weckmann M, Drömann D, Franzen KF. Impact of Heated Tobacco Products, E-Cigarettes, and Cigarettes on Inflammation and Endothelial Dysfunction. Int J Mol Sci. 2023;24. doi: 10.3390/ijms24119432

46. Ruan GT, Xie HL, Zhang HY, Liu CA, Ge YZ, Zhang Q, Wang ZW, Zhang X, Tang M, Song MM, et al. A Novel Inflammation and Insulin Resistance Related Indicator to Predict the Survival of Patients With Cancer. Front Endocrinol (Lausanne). 2022;13:905266. doi: 10.3389/fendo.2022.905266

47. Joffre J, Hellman J. Oxidative Stress and Endothelial Dysfunction in Sepsis and Acute Inflammation. Antioxid Redox Signal. 2021;35:1291–1307. doi: 10.1089/ars.2021.0027

48. Ormazabal V, Nair S, Elfeky O, Aguayo C, Salomon C, Zuñiga FA. Association between insulin resistance and the development of cardiovascular disease. Cardiovasc Diabetol. 2018;17:122. doi: 10.1186/s12933-018-0762-4

